# Leveraging High-Throughput Data to estimate Leukocyte Telomere Length in the Million Veteran Program

**DOI:** 10.1101/2025.01.14.25320560

**Authors:** Kruthika R. Iyer, Prathima Vembu, Alexa Barad, Rodrigo Guarischi-Sousa, Joseph Sarro, Paul Billing-Ross, Themistocles L. Assimes, Shoa L. Clarke, Catherine Tcheandjieu, Jennifer E Huffman, Phil Tsao, VAs Million Veteran Program

## Abstract

Telomeres are highly repeated DNA sequences that stop chromosome ends from fraying. They shorten as we age, making them markers of biological aging. In this study, we employed an innovative approach to estimate leukocyte telomere length (LTL) through the utilization of the bioinformatics tool TelSeq, leveraging existing whole-genome sequencing data sourced from the Department of Veteran Affairs’ Million Veteran Program (MVP) encompassing a cohort of 102,646 diverse individuals.

Using the TelSeq-estimated LTL, we conducted parametric and non-parametric statistical analyses with demographic descriptors such as age, sex, and genetically inferred ancestry (GIA). Our results revealed a near-linear yet modest inverse correlation between age and LTL (ρ= −0.20, p < 1E-300). Specifically, each year increase in age corresponded to an 8bp LTL decrease (p < 1E-300) after adjusting for sex and GIA groups. Females had longer LTL than males, even after controlling for ancestry and age (β = 58 bp, p < 1E-300). Additionally, differences in LTL were observed among ancestry groups (Median LTL_African_ = 2.03kb, Median LTL_European_ = 2.01kb, Median LTL_AdmixedAmerican_ = 2.00kb), however, these differences were not statistically significant between the two largest groups - African and European ancestry. To validate TelSeq-estimated LTL, we utilized methylation profiles of 140 cytosine-phosphate-guanine dinucleotides (CpGs) on 28,669 individuals to derive DNA methylation-based telomere length estimator (DNAmTL). DNAmTL had a modest correlation with TelSeq-estimated LTL (ρ=0.21, p < 1E-300). Consistent with Telseq-estimated LTL, DNAmTL was inversely associated with age where each year increase corresponded to 16bp DNAmTL decrease (p < 1E-300) after adjusting for sex and GIA groups. After controlling for these parameters, males were also found to exhibit shorter DNAmTL than females (β = −98 bp, p < 1E-300), similar to observations from TelSeq-estimated LTL. Lastly, individuals of African and Admixed-American ancestry had longer DNAmTL than those of European ancestry.

These findings align with existing literature elucidating the effects of age, sex, and ancestry on telomere length variations, and also represent the first comparison of TelSeq-derived LTL with methylation-derived DNAmTL. This valuable resource is now accessible to the broader MVP community, facilitating further telomere research.

## Introduction

Aging is a progressive functional decline driven by molecular changes and environmental factors increasing the risk of conditions such as frailty, cancer, and cardiovascular diseases^1^. With the U.S. population rapidly aging, and projections indicating older adults (>65 years) outnumbering children by 2034 and doubling by 2060^2^, accurate assessment of aging is crucial. While calendar age is closely associated with biological functioning of mammals, it often diverges from biological age due to the heterogeneity and complexity of the aging process^3,4^. To date, 12 hallmarks^5^ of biological aging have been proposed, encompassing molecular, cellular, and pathophysiological pathways. Among these, telomere length and the epigenetic clock are the two most frequently studied biomarkers, providing valuable insights into the underlying molecular mechanisms of aging.

Telomeres are guanine rich DNA-protein structures comprised of repetitive hexameric sequences (TTAGGG_n_) that cap the ends of all linear chromosomes in humans^6^. These ‘aglets’ on our chromosomes play a crucial role in preserving genetic information by protecting it from nucleolytic degradation, random recombination, repair and inter-chromosomal fusion^7^. As a part of the natural cell replication process, telomere length progressively shortens with each mitotic replication, making it a function of cell division and ostensibly a marker of biological aging^6,7^.

Maintaining optimal telomere length is crucial to overall health as both attrition and elongation may influence the risk of disease^8^. Longer telomeres have been associated with an increased risk of various cancers such as brain, lung, ovarian, bladder, neuroendocrine and skin cancer^8-10^. Conversely, telomere attenuation has been associated with several age-related conditions, including cardiovascular disease^8,11,12^, type 2 diabetes^13,14^, musculoskeletal frailty^15,16^ as well as psychiatric^17,18^, and neurological disorders^19,20^. These results, compounded by the ancestral differences observed in telomere length^21-23^, emphasize the need to investigate factors influencing telomere dynamics and homeostasis.

In the past, numerous studies have utilized laboratory assays to measure telomere length^24-35^. However, these methods incur significant expenses, demanding considerable investment of both time and resources. Advances in high-throughput sequencing technology have fostered several bioinformatic approaches that quantify telomere length by extracting telomere motif copies from whole genome sequencing (WGS) data^36-41^. Comparison of these approaches against various lab assays were previously executed by the National Heart Lung and Blood Institute’s Trans-Omics for Precision Medicine (TOPMed) program^42^. They found the TelSeq^36^ algorithm to be the best bioinformatic tool for telomere length estimation due to its computational efficiency and strong correlations with the gold standard, Southern blot^24^, and as well as the widely used technique in clinical settings, flowFISH^30,43^. More recently, the UK Biobank and the All of Us Research Program also utilized TelSeq to estimate telomere length in 462,675^44^ and 242,494^45^ samples respectively, finding it to be moderately correlated with qPCR-based telomere length estimates.

Epigenetic modifications, specifically accumulation of DNA methylation have been closely linked with the aging process^46^. Computational methods like DNAmTL^47^, which profiles genome-wide DNA methylation in CpG sites, have been developed to predict telomere length. This method was found to be moderately correlated with traditional lab assays like qPCR and flowFISH^48^.

However, to our knowledge, a direct comparison of DNAmTL with TelSeq has not been conducted. Given the tremendous surge in the availability of high-throughput data from large epidemiology studies and biobank scale research, the value of adopting bioinformatic algorithms to generate high-quality telomere length calls from WGS and methylation data cannot be understated.

At 1 million Veterans and counting, the US Department of Veterans Affairs (VA) Million Veteran Program (MVP) stands as a comprehensive biobank with a rich repository of phenomic and genomic information ^49^. In this study, we generated a new aging phenotype by utilizing WGS data obtained from 102,646 diverse MVP participants to estimate leukocyte telomere length (LTL) using the TelSeq methodology. We examined the relationship between estimated LTL and age, sex, and genetically inferred ancestry (GIA), aiming to characterize the behavior of this biomarker of aging. To validate TelSeq-derived LTL, we also leveraged methylation data from 28,669 individuals to estimate DNAmTL, evaluated TelSeq’s performance in comparison to DNAmTL before disseminating the phenotype of aging to the wider MVP scientific community.

## Methods

### MVP study population

The MVP is a nationwide multi-ancestry cohort initiated in 2011 with the objective of delineating the impacts of genetics and lifestyle factors on the health and disease outcomes of US Veterans. Blood biospecimens were acquired for the purpose of DNA extraction, genotyping, and sequencing, with age recorded at the time of the blood draw. A detailed description of the study design, recruitment, and initial characterization for this cohort has been previously published^49^.

Samples selected for WGS analysis included both a nested case-control design and random sampling [Supp Figure 1a] in order to capture disease areas of special interest to the VA but also to be broadly useful for analysis across the disease spectrum. 13,252 post-traumatic stress disorder (PTSD) cases and matched controls were included as well as 5,826 cases of chronic kidney disease (CKD), 6,001 cases of peripheral artery disease (PAD), and 340 cases of extreme hypertension (HTN). CKD, PAD, and HTN cases were oversampled for participants of African ancestry compared to MVP as a whole. A set of 29,703 common controls for the CKD, PAD, and HTN cases were selected matched on age (in 5 year bins), biological sex, self-reported race and ethnicity, smoking status, and diabetes status. In order to maximize broad cases status, and to capture healthy aging, everyone over the age of 80 at enrollment was also selected (N~16,548). Initial investigations showed that this set of 55,122 participants could be used to select appropriate case and control sets for a variety of analyses across the disease spectrum.

Subsequent sample selection included 2 participants from our metabolomics pilot and 36,746 random samples. As WGS analysis is on-going, random sampling will be used.

Methylation samples were selected based on the same phenotype criteria used for WGS. Targeted sampling was conducted on 22,383 participants to capture cases of PTSD, CKD, PAD and extreme HTN, with an oversampling of participants of African ancestry. An additional 5,104 participants aged over 80 years at enrollment were oversampled to capture healthy aging. And lastly, random sampling was performed on 17,964 participants to ensure broader representation.

### MVP whole genome sequencing and genome quality control

WGS was performed to an average depth of 30X using DNA isolated from blood, PCR-free library construction, and Illumina NovaSeq 6000 technology with a 150 bp paired end reads. Sequencing was performed by Personalis Inc. Details on variant calling through the GATK pipeline have been previously documented^50^.

Individual genome quality was evaluated using a suite of bioinformatic tools. FastQC (version 0.11.4) was employed to assess base sequence quality and GC content, with thresholds set at an average quality score of ≥28 and GC content between 40% and 41%. Read alignment quality was evaluated using Samtools (version 0.1.19), retaining only mapped paired reads with an alignment rate of ≥95%. Lastly, contamination rate was assessed using verifyBAMID, with a threshold of 5% set to exclude any genome exhibiting a higher contamination rate. These quality control procedures resulted in a sample size of 104,923 for MVP WGS Release 2.

### Estimating leukocyte telomere length (LTL) for MVP individuals with WGS data

LTL was estimated for samples with sequenced data from the MVP WGS Release 2 [Supp Figure 1b]. The phenotype was derived from raw WGS CRAM data using the TelSeq algorithm, which quantifies reads containing a fixed number of contiguous repeats of the telomere-identifying motif (TTAGGG_n_) [Figure 1]. This algorithm necessitates two primary input parameters: read length and repeat number threshold. The read length was obtained from the MVP documentation and cross-validated using the SEQ field within the analyzed CRAM files. For a read length of 150 bp, we opted a fixed repeat threshold of 12. Considering the potential for GC bias in Illumina sequencing data, where genomic regions with higher GC content affect the coverage of reads across those regions^51,52^, and recognizing that 50% of the telomeric motif comprises of GC nucleotides, rendering the telomeric region GC-rich, we applied a normalization step in TelSeq. This process adjusted for potential technical artifacts associated with GC composition by normalizing the telomeric read counts for each individual, ensuring that the GC content of the telomeric reads was between 48% and 52%.

**Figure 1:**
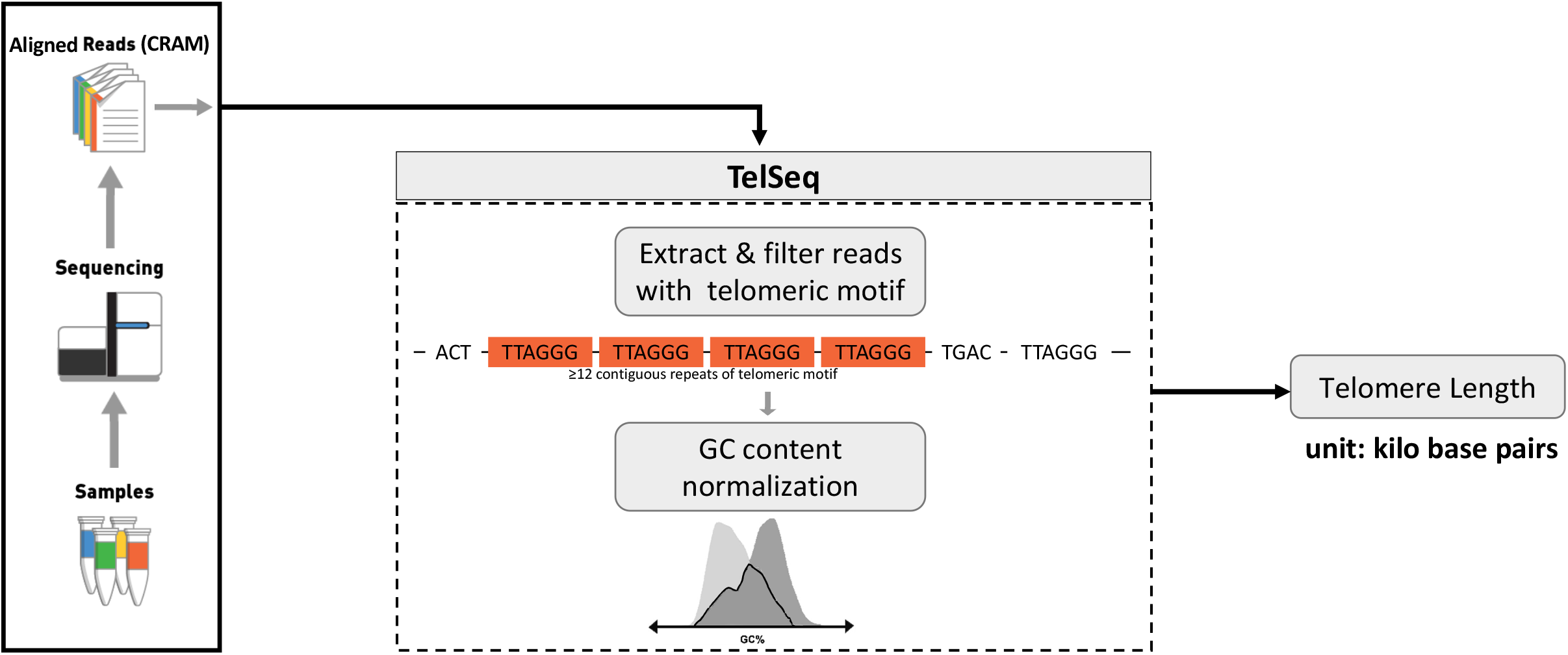
Schematic representation illustrating TelSeq workflow for the estimation of leukocyte telomere length

For the LTL estimation in MVP, we deployed TelSeq and its dependencies – bamtools and GCC, within a Docker container on the Google Cloud Platform (GCP). To facilitate the execution of TelSeq scripts on GCP, we employed the dsub job scheduler. These computations were carried out on a computing instance of the n1-standard-1 machine type, providing the compute engine with 3.75 GB of memory and 1 virtual CPU core.

### MVP methylation data, quality control and DNAmTL calculation

Methylation profiling was performed using Illumina Infinium Methylation EPIC BeadChip array v1, which includes over 850,000 probes across the human genome. Probe-level quality control consisted of excluding non-CpG probes, probes with poor mapping quality, probes exhibiting weak titration correlation or color channel switching, and those that failed the P-value with Out Of Band probes for Array Hybridization (pOOBAH)^53^ detection p value in ≥ 10% of samples. Only samples with a probe pass rate ≥ 96% were retained, resulting in a dataset of 45,460 samples with 768,569 high-quality CpG probes for MVP Methylation Data Release 1, provided in HDF5 file format. Of these samples, 28,669 had overlapping WGS data and LTL estimations [Supp Figure 1c]. Both the methylation and WGS data were obtained from the same biological sample for each individual. DNA methylation-based telomere length (DNAmTL) was estimated using a previously developed tool, which employs 140 CpGs selected via elastic net regression. Of these 140 CpGs, 113 were available in the MVP methylation data, while the remaining 27 were imputed. DNAmTL estimations were computed on the centralized MVP high-performance computing cluster using LSF job scheduler and R.

### Genetically inferred ancestry (GIA) determination within MVP

To estimate genetic ancestry, we obtained a reference dataset from the 1000 Genomes Project and used the smartpca module in the EIGENSOFT^54^ package to project the PCA loadings from a group of unrelated individuals in the reference dataset. We merged this dataset with the MVP dataset and ran smartpca to project the PCA loadings from the reference dataset. We trained a random forest classifier using continental superpopulation meta-data based on the top 10 principal components from the reference training data to define genetically inferred ancestry. We then applied this random forest to the predicted MVP PCA data and assigned ancestries to individuals with a probability greater than 50%. Those who could not be assigned to one of the five 1000G superpopulations (African (AFR), Admixed American (AMR), East Asian (EAS), European (EUR), or South Asian (SAS)) were designated “Undetermined”.

### Inferential analysis of LTL

LTL results were imported from GCP into Python computational notebooks (version 3.10.4) and integrated with age, biological sex, and GIA estimates. Stringent quality control measures were implemented, including the removal of duplicate records and the exclusion of individuals lacking GIA data, resulting in an sample size of 102,646. Due to the skewed samples sizes in certain GIA groups, coupled with deliberate oversampling of individuals over 80 years of age, our primary analysis was limited to EUR, AFR and AMR ancestry groups representing at least 5% of the overall sample size and under 80 years of age, yielding in an analytic sample size of 85,297. Subsequently, correlation analysis was conducted to elucidate the relationship between age and LTL. This analysis was further stratified by sex and GIA, allowing for a nuanced exploration of LTL variations across sexes and genetic ancestry groups. Since LTL was not normally distributed and females had small a sample size, we performed non-parametric tests to evaluate if the observed differences in LTL across these groups were statistically significant. Multi-variable linear regression was used to estimate association between LTL and age, sex, and GIA groups. Furthermore, a cubic spline model was applied to gain insight into the nature of association between age and LTL.

For primary validation analysis, we utilized a sample of 22,156 individuals representing EUR, AFR, and AMR ancestry groups, all aged ≤80 years. Correlations were conducted between TelSeq-derived LTL and DNAmTL, as well as between DNAmTL and age. Multivariable linear regression was employed to assess the association of DNAmTL with age, sex, and GIA groups. Additionally, regression models were performed with age as the outcome variable, using TelSeq-LTL and DNAmTL as predictors to evaluate their respective contributions after controlling for sex and GIA. A Bland-Altman plot was also generated to compare DNAmTL and TelSeq-derived LTL, providing insights into their agreement and potential biases.

For these analyses, the following Python and R packages were used: google.cloud.storage (version 2.7.0), pandas (version 1.5.3), matplotlib (version 3.7.1), seaborn (version 0.12.2), statsmodels (version 0.13.5), impute (version 1.80.0, Bioconductor version 3.20), and methylclock (version 1.12.0, Bioconductor version 3.20).

## Results

We analyzed 85,297 individuals from diverse genetic ancestries in MVP. The mean age was 62 years (SD = 11 years), 93% of the cohort was male and the study’s ancestry distribution was 69% European, 23% African, and 6% Admixed Americans, as determined by GIA [Table 1]. Of note, age is not normally distributed within MVP, with peaks occurring around major conflicts involving the US military as additional Americans volunteered to serve. Additionally, ~15% of the WGS samples are over age 80 due to our WGS study design criteria, compared to only 9% of all genotyped MVP participants. [Supp Figure 1a, Supp Figure 2]

For each individual within this cohort, LTL was estimated using TelSeq [Figure 1], and this process required an average time of 100 mins/participant, and resulting in a total compute time of 144611.3 hours [Supp Figure 3]. The mean LTL across these individuals was 2.05 kb (SD = 0.46 kb), spanning from 0.5 kb to 5.02 kb.

Across MVP individuals, LTL displayed a moderate negative correlation with age (ρ = −0.19; p < 1E-300). The phenotype showed statistically significant differences between the sexes (mean_male_ = 2.04kb; mean_female_ = 2.18kb; p_t-test_ = 4.74E-113; p_Mann-WhitneyUtest_ = 6.27E-85) while their correlations with age were found to be almost consistent (ρ_male_ = −0.18, p_male_ < 1E-300; ρ_female_ = −0.22, p_male_ = 1.09E-63) [Supp Figure 4].

Sample distribution was found to non-normal among GIA groups [Figure 2]. Hence a non-parametric Kruskal Wallis test was run which revealed that LTL significantly differed among the GIA-designated populations (p_KrusalWallis_ = 1.48E-10). Specifically, the mean LTL was observed to be the longest among individuals of African ancestry (mean_AFR_ = 2.07kb, IQR_AFR_ = 1.74kb – 2.36kb), followed by individuals of European ancestry (Mean_EUR_ = 2.05kb, IQR_EUR_ = 1.73kb - 2.33kb), and then Admixed Americans (mean_AMR_ = 2.00kb, IQR_AMR_ = 1.65kb – 2.37kb). The magnitude of inverse correlation with age was found to be consistent across all the ancestry groups (ρ_EUR_ = −0.20, p_EUR_ < 1E-300; ρ_AFR_ = −0.20, p_AFR_ = 9.99E-217; ρ_AMR_ = −0.16, p_HISP_ = 6.82E-38).

**Figure 2:**
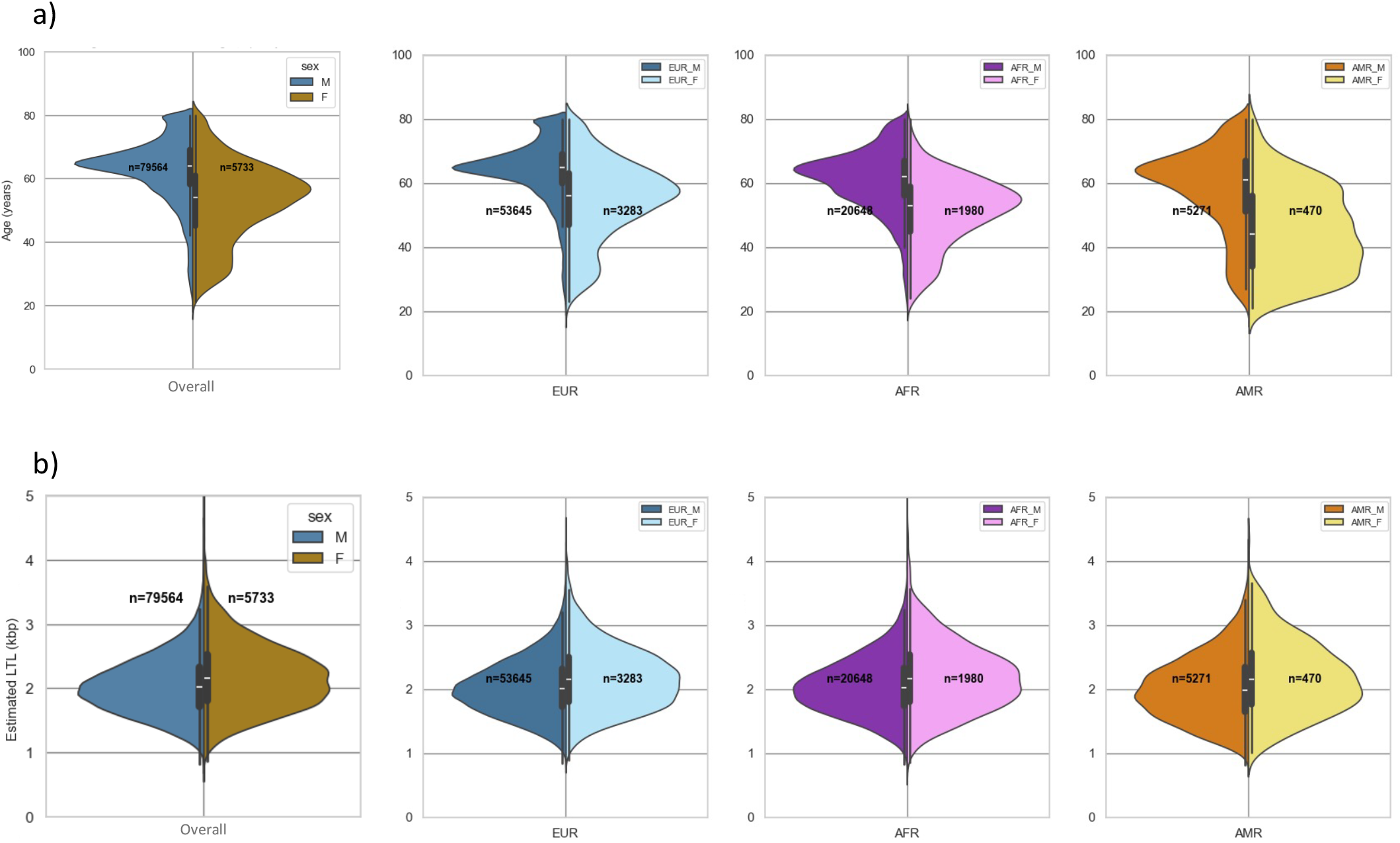
Distribution of (a) age and (b) LTL, stratified by sex and genetically inferred ancestry groups M = Males; F = Females; LTL = TelSeq estimated leukocyte telomere length in kb; AFR=African ancestry; AMR = Admixed American ancestry; EUR= European ancestry

Results of the linear regression models with LTL as the dependent variable are shown in Table 2. Considering age, sex, and ancestry groups as predictor variables, a significant inverse association was observed between age and LTL where each year’s increase in age was associated with an 8 bp (p < 1E-300) decrease in estimated LTL. Distinct sex-related differences were evident, with males exhibiting a decrease in LTL by 57 bp compared to women (p < 1E-300). With European ancestry as the reference, both Africans (β = −6 bp) and Admixed Americans (β = −8 bp) exhibited shorter LTL, with significant differences in the trend noted only in Admixed Americans (p < 1E-300).

Considering the wide age spectrum represented in our MVP cohort, we stratified the participants into 20-year age intervals and investigated the correlation between LTL and age within each specific age bracket. Consistent correlation patterns were observed in most age intervals across all ancestry groups [Supp Figure 5]. To further explore the relationship between LTL and age, we employed cubic-spline models stratified by GIA groups, while accounting for sex and age with knots at 40, 60, and 80 years. As illustrated in Figure 3, the relationship between age and LTL appeared nearly linear. This observation was reinforced by the likelihood ratio test conducted within each ancestry group, indicating that the GIA-stratified multi-variable linear model, adjusted for age and sex, provided a superior fit compared to the spline model especially in African ancestry group (p = 1.0).

**Figure 3:**
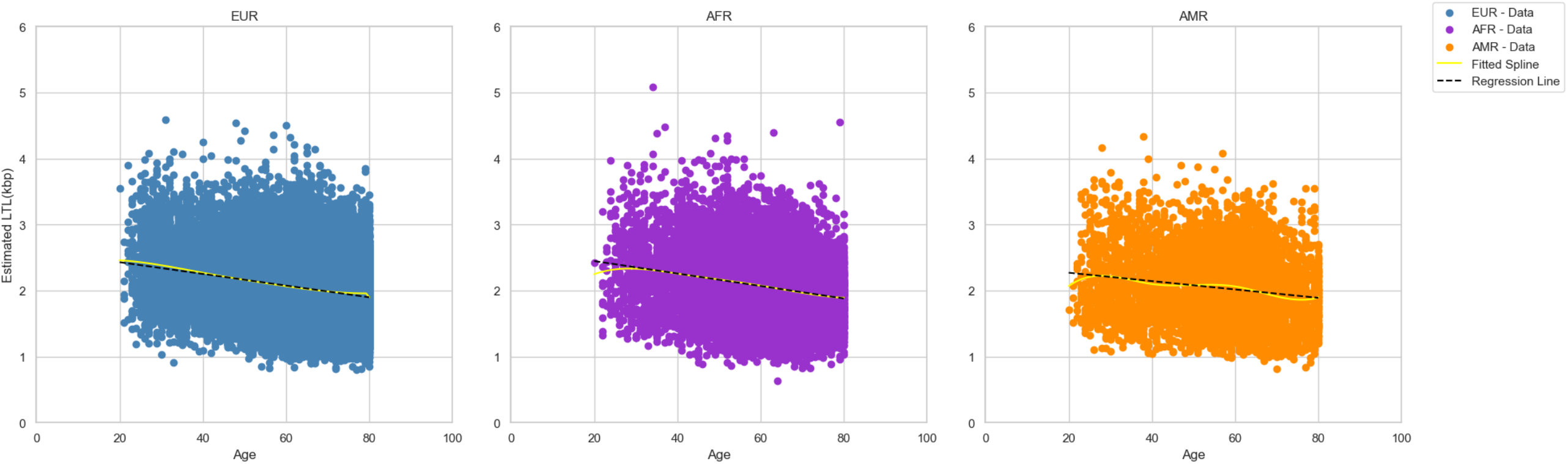
Nature of relationship between age and LTL stratified by genetically inferred ancestry groups AFR=African ancestry; AMR = Admixed American ancestry; EUR= European ancestry; LTL = TelSeq estimated leukocyte telomere length in kb

For our validation tests, which included 22,156 analytical samples, DNAmTL was positively correlated with LTL (ρ = 0.21; p < 1E-300) [Figure 4] and negatively correlated with age (ρ = −0.76; p < 1E-300). These correlations were consistent across sex and GIA groups [Figure 6-8]. Linear regression models, with DNAmTL as the outcome variable, revealed a significant inverse association with age (β = −16 bp, p < 1E-300) and male sex (β = −98 bp, p < 1E-300). Statistically significant differences in DNAmTL were observed across ancestries with African (β = 146 bp, p < 1E-300) and Admixed Americans (β = 38 bp, p < 1E-300) groups showing longer DNAmTL compared to European ancestry [Table 2]. Bland-Altman plots were also generated to assess the agreement between LTL and DNAmTL, revealing no specific trend except for a systematic difference, with DNAmTL generally estimating higher values than LTL [Figure 4]. Despite this, the vertical spread suggests that the difference between the two methods is consistent, as most samples fall within the 95% agreement limits. The absence of horizontal spread can be attributed to the sampling process. Finally, to evaluate the contributions of LTL and DNAmTL to age, we performed a regression analysis with age as the dependent variable and LTL and DNAmTL as independent predictors, adjusting for sex and GIA [Table 2]. As anticipated, both measures exhibited statistically significant inverse associations with age (β_LTL_ = −0.3 yrs, p = 0.005; β_DNAmTL_ = −28.5 yrs, p < 1E-300). The skewed male-to-female ratio in the MVP cohort, combined with the tendency for males to be older, contributed to the observed age differences by sex (β = 1.8 yrs, p < 1E-300). Additionally, the sampling strategy employed in the study underscores the age disparities observed across ancestry groups.

**Figure 4:**
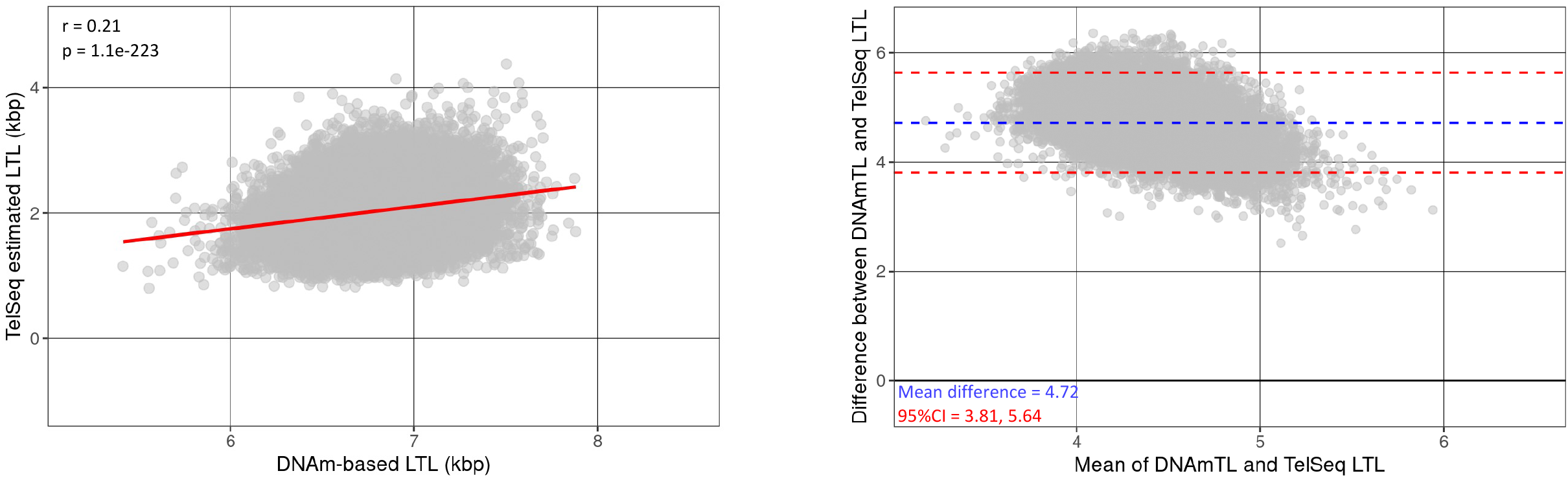
Comparison of LTL and DNAmTL using (a) Correlation Analysis (b) Bland-Altman plot to assess agreement LTL = TelSeq estimated leukocyte telomere length in kb; DNAmTL = LTL = DNA methylation estimated telomere length in kb

## Discussions

Since the Nobel Prize was awarded for the discovery of telomeres and the elucidation of their age-related shortening^55^, the field of telomere epidemiology has significantly expanded to gain deeper insights into age-related health issues and to identify potential targets for cancer therapy. Concurrently, with the decreasing cost of sequencing, there has been tremendous increase in the production of WGS data^56^, particularly from existing genetic epidemiology cohorts, through efforts such as TOPMed and the Centers for Common Disease Genomics (CCDG), and biobank-scale research initiatives, like UKBiobank, MVP, and the NIH’s All of Us research program. This surge offers a significant opportunity to capitalize on this ‘omic’ data for developing aging biomarkers. Computational tools like TelSeq—established as a cornerstone of telomere epidemiology by serving as both a quasi-standard for benchmarking other tools and a key resource for human studies—play a pivotal role in estimating telomere length in silico. In our study, we leveraged pre-existing WGS data and employed TelSeq to generate the aging phenotype of LTL. Additionally, we leveraged the largest available methylation dataset to estimate DNAmTL, validating our LTL estimates and providing a novel comparison between the two telomere length measures.

DNAmTL, a methylation-based proxy for telomere length, demonstrated a modest correlation with LTL. This is inline with the literature where traditional lab assays like TRF/FlowFISH/qPCR typically reported correlations between 0.41-0.56^47,48^. These studies, including ours, also observed wider limits of agreement in the Bland-Altman plots, suggesting limitations associated with DNAmTL. This discrepancy highlights the distinct and likely orthogonal aspects of aging captured by telomere length and DNA methylation, as evidenced by the contributions of independent predictors – LTL and DNAmTL to our age prediction model. While LTL reflects the mitotic clock^5-7^, a highly heritable trait, DNAmTL is more sensitive to environmental factors^5,46^. Furthermore, 139 of the 140 CpGs underlying DNAmTL are not located near telomere-related genes^47^, indicating that their association with aging is likely independent of telomere biology and reflects broader epigenetic modifications.

Telomere length exhibits an inverse correlation with age (ρ = −0.3), as evidenced by analysis across 124 cross-sectional studies^57^. The rate of telomere shortening is notably more rapid during early human development compared to adulthood^58,59^. Some studies observed the rate of telomere attrition decelerating around 70 years of age^60-62^, eventually reaching a plateau.

But several longitudinal studies involving elderly adults from Ashkenazi centenarians^63^, Latvian nonagenarians ^60^, and the Lothian Birth Cohorts of 1921 and 1936^64^, have revealed minimal variation in telomere length attrition. In our cross-sectional study, we observed trends that were consistent with existing literature, with a moderate negative correlation between age and telomere length that remained relatively stable across 20-year age bins. For our DNAmTL, which is reported to have a strong negative correlation with age (ρ = −0.6 − −0.8)^47,48^, we observed correlation trends similar to LTL across 20-year age bins, but the rate of attenuation appeared slightly more pronounced. Furthermore, our study suggested a linear relationship between LTL and age, but this observation could also be influenced by the left-skewed age distribution in our data, introducing potential survival bias.

In epidemiological studies, variations in telomere length have also been attributed to ancestry. The impact of genetic ancestry on telomere length garnered significant attention, with certain studies indicating that individuals of African ancestry tend to have longer telomeres compared to those of European ancestry^21-23,65^. In specific investigations, differences in average telomere length during adulthood have been attributed to longer initial telomere length among newborns of African ancestry compared to newborns of European ancestry^66-70^. Our findings revealed differences in LTL and DNAmTL based on ancestry, with individuals of African ancestry exhibited longer LTL and DNAmTL compared to those of European ancestry. It is also plausible that these differences could likely be influenced by the differential age distribution between GIA groups and rather than solely genetic ancestry. Non-European veterans predominantly served in the more recent Gulf War, Afghanistan War, and Iraq War, therefore are younger and would be expected to have longer LTL on average. This was shown by our stratified analyses where LTL and DNAmTL trends were comparable across ancestry group within age bins.

We also noted variations in LTL and DNAmTL between males and females. Although this gap may stem from the average age difference between male and female Veterans in our study (females being younger), it is established that telomere dynamics exhibit gender dimorphism, wherein adult females tend to maintain longer telomere lengths than adult males^71,72^. This sex gap has been attributed to various factors, including the protective effects of estrogen^73^, X chromosome^74^, and the hormonal milieu during fetal development^72,75^.

While our efforts represent a pioneering endeavor with the strengths of this study underscoring both diversity and large sample sizes, we acknowledge several limitations associated with the measurement technique we employed. Despite minimizing technical artifacts by using a single laboratory for DNA sample storage and extraction, and one center for sequencing and variant calling, variations in TelSeq-estimated telomere length may still arise from various sequencing-related factors: (1) telomere repeats are localized within regions of the genome that are unmapped in our sequenced CRAM files^76^, particularly in repetitive genomic regions where quality control measures may exhibit reduced stringency^77^. As a result, TelSeq may inadvertently introduce interstitial telomeric sequences —tandem repeats resembling canonical telomeric motifs located proximal to centromeres and within interstitial sites— thereby influencing the LTL estimation^37^; and (2) telomere length exhibits heterogeneity across chromosomes^78^; however, due to the inherent challenges of short-read assembly in distinguishing them on individual chromosomes, primarily because of the repetitive nature of the telomeric motif^77^, TelSeq only estimates an average LTL across all chromosomes.

Additionally, TelSeq algorithm is hardcoded to take in a fixed number of 22 pairs of autosomes into account when estimating telomere length^39^, making no adjustments for aneuploidies. For DNAmTL, it likely represents age-related changes in DNA methylation rather than directly reflecting actual telomere length, and the kilobase (kb) units retained by the algorithm can be misleading^47^, especially when comparing estimates.

In conclusion, this study marks a significant advancement by leveraging the high-throughput resources of the MVP program to bioinformatically derive a biological aging phenotype from WGS data. To our knowledge, it is also the first to compare this phenotype with an alternative estimation method utilizing the largest available methylation dataset. While LTL and DNAmTL capture distinct dimensions of aging, their integration presents an opportunity to develop a composite aging phenotype, offering a broader and more comprehensive perspective.

However, the limited overlap between the methylation and sequencing datasets constrains the composite phenotype to a smaller sample size, reducing the statistical power of subsequent analyses. Addressing this limitation will require larger methylation datasets and expanded whole-genome sequencing preferably using long-read technologies. Despite these challenges, the availability of phenotypes like LTL and DNAmTL enables researchers to investigate the genetic and clinical implications of cellular aging in a diverse population, catalyzing further biological aging research across the MVP enterprise.

## Data Availability

All data produced in the present study are available upon reasonable request to the authors

## Abbreviations

DNAmTL: DNA methylation derived telomere length
GCP: Google Cloud Platform
GCS: Google Cloud Storage
LTL: TelSeq derived Leukocyte telomere length
MVP: Million Veteran Program
VA: Department of Veterans Affairs
WGS: Whole genome sequencing

## Code availability

All custom scripts have been uploaded to a publicly accessible GitHub repository.

## Analysis pipeline

The analysis notebook and accompanying figures have been uploaded to a publicly accessible GitHub repository.

## Acknowledgements

This research is based on data from the Million Veteran Program, Office of Research and Development, Veterans Health Administration, and was supported by award #000. This publication does not represent the views of the Department of Veteran Affairs or the United States Government. We thank the participants and investigators of the MVP study who made this work possible.

## TABLES

**Table 1:** Study Demographics for MVP Telomere Length Study and All Genotyped MVP Participants N = sample size, SD = standard deviation; LTL = TelSeq eestimated leukocyte telomere length (kb); DNAmTL = DNA methylation derived telomere length; GIA = genetically inferred ancestry; AFR = African ancestry; AMR = Admixed American ancestry; EUR = European ancestry

**Table 2:** Summary table for multiple regression models in MVP N = sample size, SE = standard error; LTL = TelSeq eestimated leukocyte telomere length (kb); DNAmTL = DNA methylation derived telomere length; GIA = genetically inferred ancestry; AFR = African ancestry; AMR = Admixed American ancestry; EUR = European ancestry

## Notes

### Competing Interest Statement

The authors have declared no competing interest.

